# Tonsillectomy Outcomes for Coblation versus Bipolar Diathermy Techniques in Adult Patients: A Systematic Review and Meta-Analysis

**DOI:** 10.1101/2020.09.12.20193300

**Authors:** Abdulmalik Alsaif, Mohammad Alazemi, Narvair Kahlar, Mohammad Karam, Ahmad Abul, Abdulrahman AlNaseem, Abdulredha Almuhanna, Turki Aldrees

## Abstract

**Introduction:** There is no consensus on the most superior tonsillectomy technique in adult patients. Recent trials involving coblation technique have shown promising results.

**Aim:** The study aims to compare the outcomes of coblation versus bipolar diathermy in adult patients undergoing tonsillectomy.

**Methods:** A systematic review and meta-analysis were performed as per the Preferred Reporting Items for Systematic Reviews and Meta-analyses (PRISMA) guidelines and a search of electronic information was conducted to identify all Randomised Controlled Trials (RCTs) as well as non-randomised studies comparing the outcomes of coblation versus bipolar diathermy in adult patients undergoing tonsillectomy. Reactionary haemorrhage, delayed haemorrhage and postoperative pain were primary outcome measures. Secondary outcome measures included a return to theatre, administration of analgesia, intraoperative bleeding, diet, the effect on tonsils (degree of healing of tonsillar fossae) and operation time. Fixed effects modelling was used for the analysis.

**Results:** Four RCTs and two non-randomised studies were identified enrolling a total of 1824 patients. There were no significant differences between the coblation and bipolar groups in terms of reactionary haemorrhage (Odds Ratio [OR] = 1.81, P = 0.51), delayed haemorrhage (OR = 0.72, P = 0.20) or postoperative pain by day 7 (standardised Mean Difference [MD] = -0.15, P = 0.45). For secondary outcomes, there were no differences noted in terms of intraoperative blood loss, diet and the number of cases returned to theatre. Administration of analgesia was reported to be either insignificant between the two groups or higher in the coblation group Also, the coblation group had longer operation time and greater healing effect on tonsillar tissue.

**Conclusions:** Coblation is neither a superior or inferior option when compared to bipolar diathermy used in the current clinical practice for adult patients undergoing tonsillectomy as both techniques have similar haemorrhage rates and post-operative pain whilst also lengthening the operative time in coblation.

**Highlights:**

- Coblation is neither inferior or superior compared to bipolar diathermy in adult tonsillectomy
- Administration of analgesia either has insignificant difference or higher in the coblation group.
- Coblation has a longer operative time than bipolar diathermy.

## Introduction

Tonsillectomy is one of the commonly performed procedures globally by Otolaryngologists, and continues to be the most regularly performed surgeries in adults ^1^. Adult Tonsillectomy is indicated in chronic recurrent tonsillitis, suspicion of tonsillar malignancy, and in upper airway obstruction secondary to tonsillar hypertrophy ^2^. Over the years, various techniques and instruments have evolved to aid tonsillectomy such as bipolar diathermy dissection, harmonic ultrasonic, laser dissection and more recently the coblation method ^3^.

Coblation involves the breaking of intracellular bonds at a temperature of 60 degrees and in effect melts down the tissue^5^. Regardless of the technique used, differences are present in complications including postoperative pain, haemorrhage and postoperative infection. Hence, the debate about the optimal surgical technique continues in literature ^4^.

Diathermy involves the use of high radiofrequency current that is used to cut through or coagulate tissue ^5^. Bipolar Diathermy involves current that is being passed through two tips of the same forceps, hence the term bipolar^6^. Traditionally, diathermy was mainly used to seal the blood vessels after the surgical removal of the tonsils by a blade ^7^. The use of Bipolar Diathermy has increased in popularity ^8^ as a standalone procedure, to perform both the dissection of the tonsils and the sealing of the blood vessels to stop the bleeding. In the context of ENT, bipolar Diathermy is not only used in tonsillectomy, but it is also used to treat epistaxis. This is due to the sealing effect the heat has on the blood vessels to achieve haemostasis ^9-10^.

There is a growing amount of research around Coblation and Bipolar diathermy techniques ^11-16^, including a previous meta-analysis comparing coblation to various tonsillectomy techniques ^17^. However, up to our knowledge, there were no meta-analyses published comparing bipolar and coblation tonsillectomy techniques. Therefore, our study aims to establish the differences in efficacy between coblation and bipolar diathermy techniques and tonsillectomy outcome among adults population.

## Methods

A systematic review and meta-analysis were conducted as per the Preferred Reporting Items for Systematic Reviews and Meta-Analyses (PRISMA) guidelines ^18^.

### Eligibility criteria

All randomized control trials and observational studies comparing coblation versus bipolar diathermy haemostasis techniques for tonsillectomy were included. Coblation was the intervention group of interest and bipolar diathermy was the comparator. The study also included patients having bipolar scissors, bipolar forceps as well as cold steel and bipolar diathermy haemostasis as the comparator. All patients were included irrespective of gender or co-morbidity status as long as they belonged to either a study or control group. All case reports or cohort studies without a comparison group, studies not written in English as well as studies involving paediatric population were excluded. The study also excluded the following techniques: bipolar molecular resonance coagulation, tonsillotomy, unipolar or monopolar diathermy, adenotonsillectomy, children tonsillectomy, and conventional or traditional tonsillectomy without explicitly stating bipolar diathermy as the primary method for haemostasis.

### Primary Outcomes

The primary outcomes are haemorrhage and post-operative pain. Haemorrhage is either reactionary (within 24 hours of the operation) or delayed (after 24 hours of the operation).

### Secondary Outcomes

The secondary outcomes included a return to theatre, administration of analgesia, intraoperative bleeding and operation time (in minutes), return to normal diet (in days), and tonsillar fossae healing,.

### Literature search strategy

Two authors AA and MK independently searched the following electronic databases: MEDLINE, EMBASE, EMCARE, CINAHL and the Cochrane Central Register of Controlled Trials (CENTRAL). The last search was run on the 12^th^ of April 2020. Thesaurus headings, search operators, and limits in each of the above databases were adapted accordingly. Beside this, World Health Organization International Clinical Trials Registry (http://apps.who.int/trialsearch/),

ClinicalTrials.gov (http://clinical-trials.gov/), and ISRCTN Register (http://www.isrctn.com/) were searched for details of ongoing and unpublished studies. The search terminologies included “coblation”, “bipolar” and “tonsillectomy”. The bibliographic lists of relevant articles were also reviewed.

### Selection of Studies

The title and abstract of articles identified from the literature searches were assessed independently by two authors. The full texts of relevant reports were retrieved and those articles that met the eligibility criteria of our review were selected. Any discrepancies in study selection were resolved by discussion between the authors.

### Data Extraction and Management

An electronic data extraction spreadsheet was created in line with Cochrane’s data collection form for intervention reviews. The spreadsheet was pilot-tested in randomly selected articles and adjusted accordingly. Our data extraction spreadsheet included study-related data (first author, year of publication, country of origin of the corresponding author, journal in which the study was published, study design, study size, the clinical condition of the study participants, type of intervention, and comparison), baseline demographics of the included populations (age and gender) and primary and secondary outcome data. The data and results were cooperatively collected and recorded and any disagreements were solved via discussion.

### Data synthesis

Data synthesis was conducted using the Review Manager 5.3 software. The extracted data were entered into Review Manager by three independent authors. The analysis involved used was based on fixed effects modelling. The results were reported in forest plots with 95% Confidence Intervals (CIs).

For dichotomous outcomes, the Odds Ratio (OR) was calculated between the 2 groups. The OR is the odds of an event in the coblation group compared with the bipolar group. An OR of greater than 1 for the reactionary haemorrhage and delayed haemorrhage would favour the coblation group, an OR of less than 1 would favour the bipolar group, and an OR of 1 would favour neither groups.

For continuous outcomes, the Mean Difference (MD) was calculated between the 2 groups. A positive MD for the post-operative pain score by day 7 would favour the coblation group, a negative MD would favour the bipolar group and an MD of 0 would favour neither groups.

### Assessment of Heterogeneity

Heterogeneity among the studies was assessed using the Cochran Q test (x2). Inconsistency was quantified by calculating I^2^ and interpreted using the following guide: 0% to 25% may represent low heterogeneity, 25% to 75% may represent moderate heterogeneity, and 75% to 100% may represent high heterogeneity.

### Methodological quality and risk of bias assessment

Two authors independently assessed the methodological quality as well as the risk of bias for articles matching the inclusion criteria. A third author was used as an adjudicator if a decision was disputed. For randomized trials, the Cochrane’s tool for evaluating the risk of bias was used. Domains assessed included selection bias, performance bias, detection bias, attrition bias, reporting bias, and other sources. RCT studies are classified as studies into low, unclear, and high risk of bias. For non-randomized studies, the Newcastle-Ottawa Scale^20^ was used. It uses a star grading system to assess studies in terms of three domains: selection, comparability, and exposure. The total maximum score for each study is 9 stars.

## Results

### Literature search results

The search strategy retrieved 546 studies and after a thorough screening of the retrieved articles using the inclusion and exclusion criteria, the authors identified 6 studies in total which met the eligibility criteria (Figure 1).

**Figure 1:**
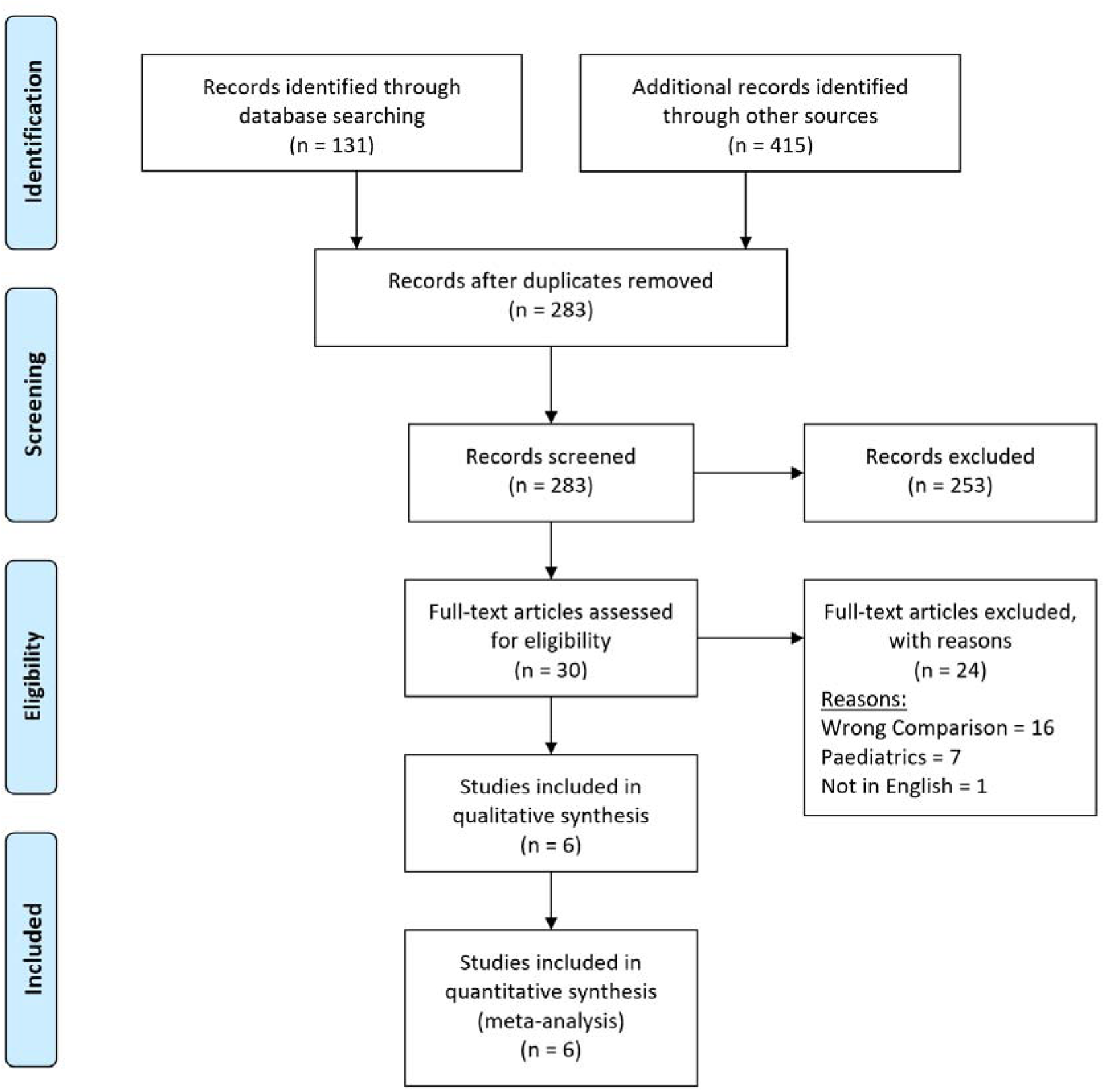
Prisma Flow Diagram. The PRISMA diagram illustrating the search and selection processes applied during the overview. PRISMA, Preferred Reporting Items for Systematic Reviews and Meta-Analyses.

### Description of Studies

#### Timms et al. 2002

Timms et al. performed single-centre double-blinded RCT that included 10 adult patients with recurrent tonsillitis undergoing routine tonsillectomy. The randomization of patients was done through storing numbers in opaque envelopes, to have one tonsil removed completely by tissue coblation, and the other by bipolar dissection. The surgical techniques on each side were not mentioned to the patients. Bipolar dissection was done using the standard technique (set at 4/10- and 50-Watts power). All tonsillectomies were extracapsular. They have excluded patients with history quinsy, medical disease, and bleeding disorders ^11^.

#### Noon et al. 2003

Noon et al. conducted a single-centre retrospective study that included 65 adult patients undergoing tonsillectomy. The study included patients undergoing coblation tonsillectomy or bipolar diathermy. At the end of this period, one researcher has analysed the patients’ notes. Bipolar dissection was done using the standard technique (set at 4/10- and 50-Watts power). All tonsillectomies were extracapsular. In addition, the study excluded obese patients.^12^

#### Polites et al. 2006

Polites et al. performed a single centred double-blinded prospective RCT that included 20 adult patients listed for tonsillectomy due to symptoms of tonsillitis. Randomly, each patient had one tonsil selected for removal by cold steel dissection with bipolar haemostasis and the other tonsil by coblation. Bipolar dissection was done using the standard technique (set at 4/10- and 50-Watts power). All tonsillectomies were extracapsular. They have excluded patients with history of peritonsillar abscess and bleeding disorders ^13^.

#### Hasan et al. 2007

Hasan et al. conducted a single centred RCT that included 40 adult patients listed for tonsillectomy due to symptoms of tonsillitis. Patients were randomly allocated into 2 groups: 20 patients underwent coblation and the other 20 were operated with bipolar scissors. The randomization was generated by using sequence number on the day of the operation. All tonsillectomies were extracapsular. They have excluded patients with history of peritonsillar abscess, major health problems and bleeding disorders.^14^

#### Belloso et al. 2010

Belloso et al. conducted a single-centre prospective observational cohort study that included 318 coblation tonsillectomies with a control group of 261 tonsillectomies performed by blunt dissection from July 2001 to January 2003. The groups reported in the study were extracted from a tonsillectomy audit. All tonsillectomies were extracapsular^15^.

#### Alvarez Palacios et al. 2017

Alvarez Palacios et al. performed single-centre RCT that included 103 adult patients who were scheduled for elective tonsillectomy. Patients were randomized to tonsillectomy with cold dissection (CD), monopolar-bipolar dissection (MBD), and coblation dissection (CBD). All tonsillectomies were extracapsular^16^.

**Table 1.** Baseline Characteristics of the Included Studies.

| Study (Year) | Country, Journal | Study Design | Sex (Male: Female) | Mean Age ± SD (Range) | Total Study Sample (Control: Intervention) | Interventions Compared |
| --- | --- | --- | --- | --- | --- | --- |
| Timms et al. (2002) | The Journal of Laryngology & Otology, UK | RCT | 3:7 | 25.3 years (18-36 years) | 10 patients (10:10 tonsils) | Coblation versus Bipolar Diathermy |
| Noon et al. (2003) | The Journal of Laryngology & Otology, UK | Retrospective Analysis | 17:48 | Coblation: 25.1 years (16-50 years)<br>Bipolar: 21.8 years (16-36 years) | 65 (36:29) | Coblation versus Bipolar Dissection |
| Polites et al. (2006) | ANZ Journal of Surgery, Australia | RCT | 10:9 | 26.3 years (16-41 years) | 19 patients (19:19 tonsils) | Cold steel dissection with bipolar haemostasis versus coblation dissection |
| Hasan et al. (2007) | European Archives of Oto-Rhino-Laryngology, Finland | RCT | 16:24 | NR (18-55 years) | 40 (20:20) | Coblation versus bipolar scissors |
| Belloso et al. (2010) | The Laryngoscope, UK | Cohort | NR | Pediatrics<br>Coblation: $7.76 \pm 3.56$ years<br>Bipolar: $7.60 \pm 3.44$ years<br>Adults<br>Coblation: $29.69 \pm 14.27$ years<br>Bipolar: $28.20 \pm 11.60$ years | Total: 1587 (844:743)<br>Paediatrics: 1008 (526:482)<br>Adults: 579 (318:261) | Coblation versus blunt dissection with bipolar diathermy hemostasis |
| **Alvarez**<br>**Palacios et al.**<br>**(2017)** | Indian Journal of<br>Otolaryngology<br>and Head & Neck<br>Surgery, Spain | RCT | Coblation<br>dissection: 12:15<br>Monobipolar<br>Diathermy: 16:25<br>Cold Dissection:<br>16:19 | 26.1 ± 7.2 years (16-50 years) | 68 (27:41) | Cold Dissection,<br>Coblation Dissection<br>and Monopolar<br>dissection + Bipolar<br>haemostasis |

### Primary Outcomes

#### Haemorrhage

Haemorrhage was divided into reactionary haemorrhage (within 24 hours post-operatively) and delayed haemorrhage (after 24 hours post-operatively).

In Figure 2, reactionary haemorrhage was reported in four studies enrolling in 193 patients. There was no statistically significant difference seen in the odd ratio analyses showing a lower rate of reactionary haemorrhage for the coblation group (OR = 1.81, CI = 0.31 to 10.40, P = 0.51). A moderate level of heterogeneity was found amongst the studies (1^2^ = 42%, P = 0.19).

**Figure 2:**
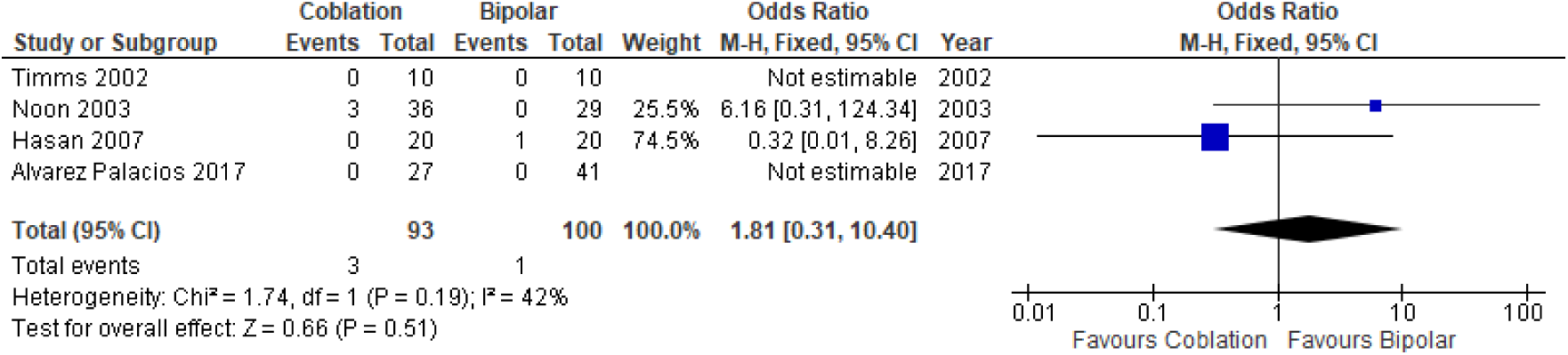
Forest Plot of Coblation versus Bipolar Tonsillectomy - Reactionary Haemorrhage. Quantitative analysis showing the odds ratio in the reactionary haemorrhage reported by Timms et al. (2002), Noon et al. (2003), Hasan et al. (2007) and Alvarez Palacios et al. (2017)

In Figure 3, delayed haemorrhage was reported in six studies enrolling 810 patients. There was no statistically significant difference seen in the odd ratio analysis illustrating a lower rate of delayed haemorrhage for the coblation group (OR = 0.72, CI = 0.43 to 1.19, P = 0.20). A moderate level of heterogeneity was found amongst the studies (I^2^ = 59%, P = 0.01).

**Figure 3:**
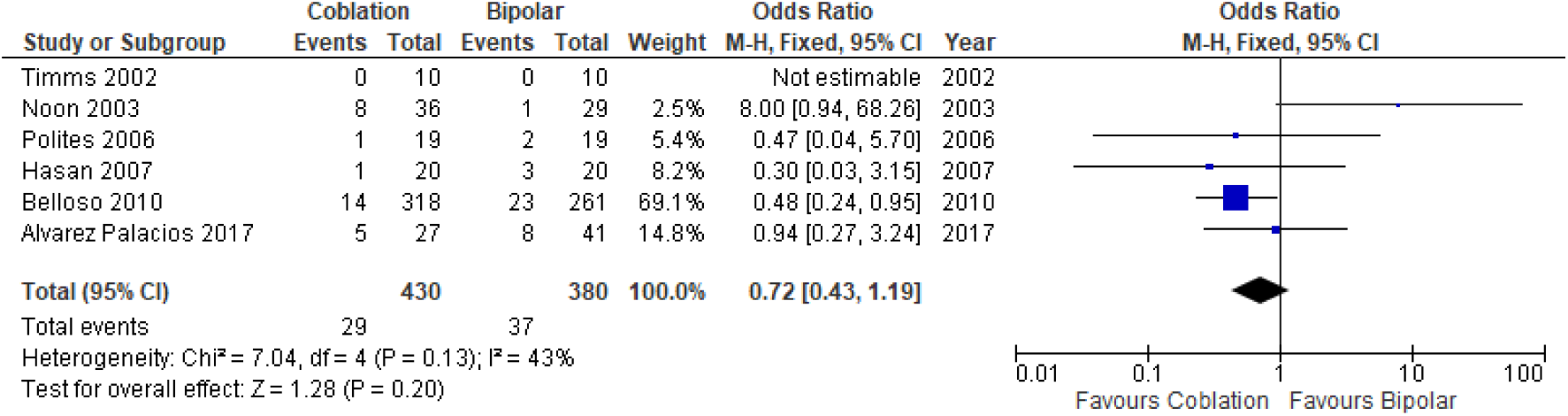
Forest Plot of Coblation versus Bipolar Tonsillectomy - Delayed Haemorrhage. Quantitative analysis showing the odds ratio in the delayed haemorrhage reported by Timms et al. (2002), Noon et al. (2003), Polites et al. (2006), Hasan et al. (2007), Belloso et al. (2010) and Alvarez Palacios et al. (2017).

#### Post-operative Pain by Day 7

In Figure 4, post-operative pain by day 7 was reported using different pain scores in two studies enrolling 106 patients. There was no statistically significant difference seen in the standardized mean difference analysis showing less post-operative pain associated with the coblation group (standardized MD = -0.15, CI = -0.54 to 0.24, P = 0.45). A low level of heterogeneity was found amongst the studies (I^2^= 0%, P = 0.64).

**Figure 4:**
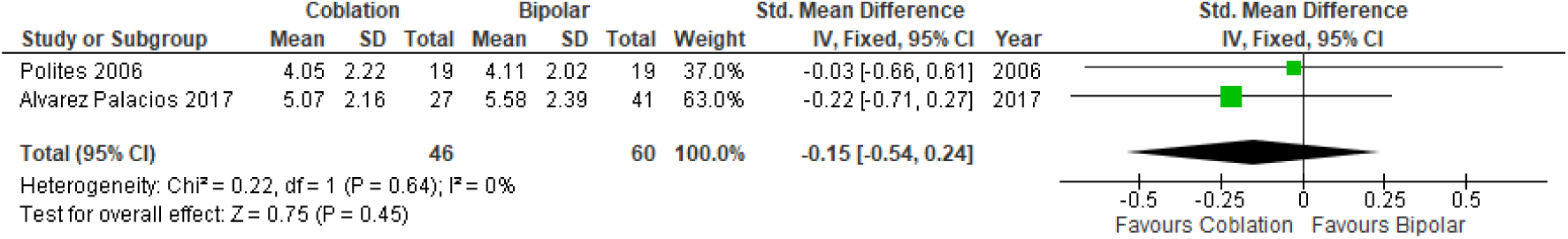
Forest Plot of Coblation versus Bipolar Tonsillectomy - Postoperative Pain by Day 7. Quantitative analysis showing the standardized mean difference in post-operative pain by day 7 reported by Polites et al. (2006) and Alvarez Palacios et al. (2017).

**Timms, 2002 and Hasan, 2007** also reported post-operative pain scores on day 7 however this was given as a median (and range) therefore not allowing quantitative analysis. Timms showed a significant median score favouring the coblation group (p = 0.001) whilst Hasan showed an insignificant difference between the two groups (p = 0.962).

### Secondary outcomes

#### Return to theatre

Noon et al reported no significant difference in the number of cases that returned to the theatre between coblation and bipolar groups (4 and 0 cases, respectively)

#### Administration of Analgesia

Hasan et al and Alvarez Palacios et al looked into the administration of analgesia following both coblation and bipolar diathermy. Hassan used a self-administered device containing opioids and showed a higher median number of analgesic doses required by the coblation group during the hospital stay compared to the bipolar group (9 versus 4, respectively). Alvarez Palacios et al used a combination of paracetamol, codeine, dexketoprofen, and Metamizole ampoles reporting no significant difference in the average consumption of analgesics during the first post-operative day.

#### Intra-operative Bleeding

Hasan et al. reported no significant difference between the coblation and bipolar groups in terms of intraoperative bleeding.

#### Diet

The time taken to return to a normal diet was reported by Hasan et al. The study found no significant difference in the patients’ opinion of their ability to return to a normal diet while comparing the two distinctive groups.

#### Effect on Tonsils: Degree of healing in tonsillar fossae

Timms et al. reported a significant difference in the healing of the tonsillar fossae between post-coblation and post-bipolar dissection groups. The results found that the post-coblation group had more rapid healing of the tonsillar fossae. In the coblation group: 7 tonsils were fully healed, one tonsil had 50% slough and two tonsils had only 25% slough. In the bipolar group: 9 tonsils were covered with 100% slough and one tonsil had 50% slough.

#### Operation time (min)

The operative time (min) between coblation and bipolar dissection groups was only reported by Hasan et al who found a significant difference between the two groups. In the coblation group, the mean time taken to operate was 20.5 minutes whilst the bipolar diathermy group had a mean operating time of 12 minutes with a p-the value of <0.001.Methodological Quality and Risk of Bias Assessment The Cochrane Collaboration tool was used to assess the quality of the RCTs included in the study (Table 2). The Newcastle-Ottawa scale was used to assess the quality of the non-randomized studies (Table 3) which offers a star system for analysis^19^. The quality of the two non-randomized studies (Belloso et al. and Noon et al.) was deemed to be high in selection and exposure but low incomparability. Overall, both studies were of good quality based on the AHRQ standards ^19^.

**Table 2.** Bia analysis of the Randomised Trials using the Cochrane Collaboration’s Tool

| First Author | Bias | Authors' Judgement | Support for Judgement |
| --- | --- | --- | --- |
| **Timms et al. (2002)** | Random sequence generation (selection bias) | Low Risk | Random envelopes were used |
|  | Allocation concealment (selection bias) | Low Risk | Numbers stored in opaque envelopes |
|  | Blinding of participants and personnel (performance bias) | Low Risk | Double-blinded RCT |
|  | Blinding of outcome assessment (detection bias) | Low Risk | Patients were seen by a different surgeon in the outpatient department who did not know the surgical technique has done |
|  | Incomplete outcome data (attrition bias) | Low Risk | No missing outcome data |
|  | Selective reporting (reporting bias) | Unclear Risk | No information is given |
|  | Other bias | Low Risk | The study appears to be free of other sources of bias. |
| **Polites et al. (2006)** | Random sequence generation (selection bias) | Unclear Risk | Randomization method not mentioned |
|  | Allocation concealment (selection bias) | Unclear | No information is given |
|  | Blinding of participants and personnel (performance bias) | Low Risk | Double-blinded RCT |
|  | Blinding of outcome assessment (detection bias) | Low Risk | Outcome measurement is not likely to be influenced |
|  | Incomplete outcome data (attrition bias) | Low Risk | No missing outcome data |
|  | Selective reporting (reporting bias) | Low Risk | All outcome data reported |
|  | Other bias | Low Risk | Similar baseline characteristics in both groups |
| **Hasan et al. (2007)** | Random sequence generation (selection bias) | Low Risk | Randomized via a number generated sequence |
|  | Allocation concealment (selection bias) | Unclear | No information is given |
|  | Blinding of participants and personnel (performance bias) | High Risk | Single-blinded RCT- the surgeon was not blinded |
|  | Blinding of outcome assessment (detection bias) | Low Risk | Single-blinded RCT- patients and nursing staff were blinded. |
|  | Incomplete outcome data (attrition bias) | Unclear Risk | No missing outcome data however 5 patients were excluded from analysis (3 returning to theatre patients, 2 not completing Post-operative pain ratings) |
|  | Selective reporting (reporting bias) | Low Risk | All outcome data reported |
|  | Other bias | Low Risk | Similar baseline characteristics in both groups |
| **Alvarez Palacios et al. (2017)** | Random sequence generation (selection bias) | Unclear Risk | Randomization method not mentioned |
|  | Allocation concealment (selection bias) | Unclear Risk | Information not mentioned |
|  | Blinding of participants and personnel (performance bias) | Unclear Risk | No information is given |
|  | Blinding of outcome assessment (detection bias) | Low Risk | Outcome measurement is not likely to be influenced |
|  | Incomplete outcome data (attrition bias) | Low Risk | No incomplete outcome data |
|  | Selective reporting (reporting bias) | Low Risk | All reported |
|  | Other bias | Low Risk | Similar baseline characteristics in both groups |

**Table 3.** Newcastle-Ottawa Scale to Assess the Quality of Non-randomised Studies.

| Study | Selection | Comparability | Exposure |
| --- | --- | --- | --- |
| **Belloso et al. (2010)** | **** | * | *** |
| **Noon et al. (2003)** | **** | * | *** |

## Discussion

Coblation showed a similar effect when compared with bipolar diathermy with all primary outcomes as shown by the results of the analyses. Reactionary haemorrhage showed no significant (P=0.51) improvements in the coblation group when compared with the bipolar group (Figure 2). Also, there were a similar number of delayed haemorrhage cases within both groups (P =0.20) (Figure 3). Furthermore, postoperative pain scores also had no significant difference (P=0.45) in both techniques (Figure 4). The between-study heterogeneity was deemed moderate for reactionary and delayed haemorrhages (I^2^ = 42 and 43% respectively), however, the heterogeneity for postoperative pain score was judged to have a low level (I^2^= 0%), based on the assessment as mentioned in Section 2.

In addition to the outcomes mentioned above, the findings of this study reported many secondary outcomes that proved coblation to has very similar efficacy to bipolar diathermy for adult patients undergoing tonsillectomy. Intraoperative blood loss and diet both showed similar results between both techniques. Similarly, there were no significant differences noted in terms of the number of cases that returned to the theatre. In addition, operative time was found to be longer in the coblation group. Furthermore, the rate of healing of the tonsillar fossae was seen to be greater in coblation compared with the bipolar technique. Regarding the administration of analgesia, Hasan et al found that the coblation group had a significantly higher number of analgesic doses during the hospital stay, while Alvarez Palacios et al found no significant difference between the two groups.

Even though tonsillectomy is one of the most commonly performed procedures in otorhinolaryngology and worldwide, none of the popularly used techniques for tonsillectomy is superior to date ^20^. The current study findings show that coblation is not a superior or an inferior option to bipolar diathermy for adult patients undergoing tonsillectomy. The finding of an insignificant difference in the rates of haemorrhage is supported by the work done by Leinbach et al. that looked into tonsillectomy techniques and found no significant difference in both groups ^21^.

However, other studies found that coblation had a higher rate of postoperative haemorrhage ^17^. This controversy in the findings was addressed by Ragab et al. who mentioned that there are not enough large studies to detect the small difference in the rates of haemorrhage ^22^. Additionally, the work by Philpott et al. found that coblation had no symptomatic advantages over conventional dissection tonsillectomy, which supports the findings of this study regarding postoperative pain ^23^. In contrast, Cardozo et al. found a significant association between bipolar diathermy and post-operative pain in adult tonsillectomy ^24^. With regards to the healing of the tonsillar fossae, Temple et al. found all fossae to be healed in patients who underwent coblation tonsillectomy, which also supports the current study’s findings, but their study only included paediatric patients ^25^.

A study conducted in the United Kingdom highlighted that coblation is a cost-effective intervention for tonsillectomy when compared with cold steel procedures for both adult and paediatric patients, with a probability of at least 0.7 at being cost-effective ^26^. In another study conducted in the United States, coblation was reported to have significantly lower costs compared to electrocautery surgery, however, the total cost of coblation was slightly higher when central supply was taken into account^27^.

Based upon the results from the best available evidence comparing coblation tonsillectomy to bipolar diathermy in adult patients, there is no significant difference in the outcomes between the two techniques apart from more rapid healing of the tonsillar fossae in the coblation technique. The current evidence, therefore, suggests coblation tonsillectomy does not necessarily need to replace bipolar diathermy in adult patients undergoing tonsillectomy. However, complete certainty is difficult due to the controversial findings of the studies previously mentioned, highlighting the importance of additional well-designed, large, randomized control trials in order to draw a more definitive conclusion.

A systematic approach was applied to provide a strong conclusion based on reliable evidence. The study included four standardized RCTs and two non-randomized studies with a total sample size of 1824 patients of a wide range of ages. The large sample size and age ranges allow the study to be more representative of the general population, providing a better perspective and understanding. However, the current study also had some limitations. The search strategy only yielded six studies which also lacked the statistical power to reach appropriate conclusions regarding the effectiveness of coblation compared to traditional tonsillectomy. Due to the nature of the treatment being studied, double-blind randomization was not possible. In addition, included studies contained heterogeneous populations in terms of ages, surgical indications, and types of procedures. Furthermore, uncontrollable variables, such as the ‘skill level’ of surgeons performing such operations may have influenced the outcomes.

## Conclusions

Although the evidence is limited with only six studies comparing coblation and bipolar diathermy, the results of this meta-analysis suggest that coblation does not improve or worsen the outcomes in adult patients undergoing tonsillectomy as it has similar haemorrhage cases and comparable postoperative pain scores, whilst also lengthening the operative time. The authors suggest further clinical studies to be performed to determine whether coblation has any superior clinical benefits.

## Data Availability

The datasets generated and analysed during the current study are available from the corresponding author on reasonable request.

## Appendix 1. Abbreviation List

| **Abbreviation** | **Stands for** |
| --- | --- |
| **RCT** | Randomised Controlled Trial |
| **CIs** | Confidence Intervals |
| **OR** | Odds Ratio |
| **MD** | Mean Difference |
| **PRISMA** | Preferred Reporting Items for Systematic Reviews and Meta-Analyses |

## Declarations

### Ethics Approval and Consent to Participate

Not Applicable.

### Consent for Publication

Not Applicable.

### Competing Interests

The author(s) declared that they have no competing interests.

### Funding

The author(s) received no financial support for the research, authorship, and/or publication of this article.

### Author Contributions

Abdulmalik Alsaif and Mohammad Alazemi contributed equally to the paper as first co-authors. Data Acquisition: Abdulmalik Alsaif, Abdulrahman AlNaseem, Abdulredha Almuhanna, Ahmad Abul and Mohammad Karam

Data Analysis and Interpretation: Abdulmalik Alsaif, Ahmad Abul and Mohammad Karam Mohammad Alazemi, Narvair Kahlar and all the other authors contributed to drafting the manuscript.

Study Concept and design: Ahmad Abul and Mohammad Karam

Study supervision and logistic support: Turki Aldrees

All authors read and approved the final manuscript.

## Acknowledgements

Not Applicable.

## Notes

### Competing Interest Statement

The authors have declared no competing interest.

